# Antimicrobial Resistance Profile of Bacterial Uropathogens Obtained from Patients in Tertiary Healthcare Facilities in Calabar, Southern Nigeria

**DOI:** 10.1101/2025.04.24.25326348

**Authors:** Emmanuel Effiong Bassey, Manal Mohammed, Edet E. Ikpi, Anyanwu A. A. Alaribe

## Abstract

**Background:** Urinary tract infections (UTIs) are major health concerns affecting millions of people globally each year, and their management is complicated by increasing resistance to antibiotics among uropathogens. Hence, this study was conducted to determine the status of multidrug resistance (MDR) among bacterial uropathogens from symptomatic patients attending tertiary healthcare facilities in Calabar, Nigeria.

**Methods:** A total of 65 uropathogens isolated from symptomatic UTI patients from tertiary healthcare facilities in Calabar were identified using standard microbiological methods. Gram-negative uropathogens were further confirmed with commercial biochemical test kits: Analytical Profile Index (API) 20E and API 20NE (Bio Mérieux). Antibiotic susceptibility of the isolates was determined according to modified Kirby–Bauer method following the Clinical Laboratory Standards Institute recommended guidelines.

**Results:** *Klebsiella pneumoniae* (23.1%) was the predominant isolate, followed by *Staphylococcus* sp., (16.9%) and *Escherichia coli* (12.3%). Other isolates recovered include *Enterobacter clocae*, *Citrobacter freundii*, *Proteus mirabilis*, *Serratia marcescens*, *Pseudomonas aeruginosa*, *Cronobacter* sp., *Enterococcus* sp., *Citrobacter koseri*, and *Pseudomonas luteola*. Most of the isolates were highly resistant to augmentin, amoxicillin, septrin, ampiclox, erythromycin, and rifampicin. However, bacterial strains were sensitive to levofloxacin, gentamicin, ciprofloxacin and sparfloxacin.

**Conclusion:** This research provides an update on the trends in bacterial pathogens causing UTIs, and MDR among uropathogens in different hospitals. This information will assist clinicians in making antibiotic choices in the management of UTIs. To reduce the emergence of resistance in bacterial uropathogens, prudent use of antimicrobial agents is advised.

## Introduction

Urinary tract infections (UTIs) are severe public health conditions characterized by the presence of pathogenic microorganisms in the urinary tract (Bassey et al., 2024; Muhammad et al., 2020). Microbial invasion can lead to inflammation of the urinary epithelium (urothelium), including the bladder (cystitis), kidney (pyelonephritis), ureter (urethritis), and urethra (urethritis) (Farag et al., 2024). Annually, 150 million people worldwide are affected by UTIs in developed and developing countries (Flores-Mireles et al., 2015). UTIs are caused mainly by gram-negative bacteria, accounting for 80–90% of cases; however, fungi, parasites, and viruses are occasionally involved in UTIs (Bitew et al., 2022; Seifu & Gebissa, 2018).

UTIs can affect individuals of all age groups, but their prevalence also increases with advancing age, catheterization, sexual promiscuity, menopause and prostrate problems (Almukhtar, 2018; Jung & Brubaker, 2019). Women are more vulnerable to UTIs, and approximately 60% of women experience UTIs in their lives. This is due to their short urinary anatomical structure, physiological changes caused by menstrual flow, changes in vaginal pH due to the depletion of commensal bacteria and pregnancy (Huang et al., 2022; Ku et al., 2024). In addition, diabetic women have an increased risk of contracting UTIs (Schneeberger et al., 2018).

The management of UTIs, especially in developing countries, is based on empirical antibiotic treatment. Unfortunately, the overuse of antibiotics in clinical settings without prior knowledge of the sensitivity patterns of causative organisms increases resistance to commonly prescribed drugs. In recent times, the prevalence of multidrug-resistant (MDR) uropathogens has increased tremendously, leading to increased risks of treatment failure, high healthcare costs and high morbidity and mortality of UTI patients (Ku et al., 2024; Alhazmi et al., 2023; Tessema et al., 2020). Although the Infectious Diseases Society of America recommends trimethoprim-sulfamethoxazole (TMP-SMX), nitrofurantoin, or fosfomycin as first-line therapy in UTI management, local knowledge of the causative agent involved in UTIs and their antibiotic susceptibility pattern is crucial for effective treatment options and prevention of the emergence of antimicrobial resistance (Muhammad et al., 2020).

In Calabar, Nigeria, UTIs are among the leading causes of hospital visits. However, effective treatment of UTIs is often hindered by poor utilization of empiric antibiotic regimens (Ogban et al., 2020), underscoring the importance of effective empirical antibiotics for the management of UTIs. The present study was conducted to determine the status of MDR among bacterial uropathogens from symptomatic patients attending tertiary healthcare facilities in Calabar, Nigeria. This research provides an update on the trends in bacterial pathogens causing UTIs, and MDR among uropathogens in different hospital settings within the study area.

## Materials and Methods

### Study area/design and population

A cross-sectional study was conducted at three tertiary healthcare facilities in Calabar, viz., the University of Calabar Teaching Hospital (UCTH), Nigerian Navy Reference Hospital (NNRH), Calabar and General Hospital Calabar (GHC), between September and December 2021 to investigate the etiology and antibiotic profile of uropathogens among patients diagnosed with UTI. These hospitals were chosen because they are major healthcare providers for both inpatients and outpatients in Cross River State, the neighboring states (Benue, Ebonyi, Abia and Akwa Ibom) and the Cameroon Republic. The sample size of 240 was determined by the Kish (1965) formula, *N* = *Z*^2^*p* (1− *p*)/*d*^2^, where; *Z* = *Z* score for 95% confidence interval = 1.96, *p* = prevalence (19.0%), and *d* = acceptable error or precision (0.05). Study participants were randomly recruited until the expected study sample was attained.

### Inclusion and exclusion criteria

Inpatients and outpatients above five years of age, those clinically diagnosed with UTIs, those whose residents are within Calabar metropolis and those receiving treatment in the health facilities under study were enrolled in the study. Patients under the age of five, those with polymicrobial infections involving more than two bacterial species, and those with a history of antibiotic therapy in the last seven days prior to the day of sample collection were excluded from the study.

### Sample collection, processing and identification of bacteria

Prior to the collection of urine samples, patients were screened to determine whether they had signs and symptoms of UTI, such as fever, urgency, frequent urination, flank or suprapubic pain, and dysuria. Clean catch midstream urine (MSU) was collected into sterile clinical urine bottles clearly labeled with the patient’s laboratory number and the assigned hospital code. In patients with urinary catheters, urine samples were collected through the catheter valve into sterile urine containers. Each batch of samples was kept in a cold box and conveyed within two hours of collection to the Bacteriology Laboratory of General Hospital Calabar for analysis.

Each urine sample was inoculated on MacConkey agar and Cystine Lactose Electrolyte Deficient (CLED) agar plates using a calibrated loop (0.002 mL) and then incubated aerobically at 37°C for 18–24 hours. Discrete colonies from significant cultures were sub-cultured onto fresh agar plates and stored in nutrient agar slants. The isolated uropathogens were identified using standard microbiological techniques, i.e., growth characteristics, Gram stain reactions, and conventional biochemical tests (Chessbrough, 2006). Gram-negative uropathogens were further confirmed with commercial biochemical test kits; the Analytical Profile Index (API) 20E and API 20NE (Bio Mérieux).

### Antimicrobial susceptibility profile of uropathogens

The uropathogens were tested against antibiotics using the disc diffusion method on Mueller‒Hinton agar (MHA) (Bassey et al., 2022). The test organisms were standardized to match 0.5 MacFarland standard. The inoculum was streaked onto the surfaces of MHA plates using a sterile cotton swab to ensure lawn growth following incubation. The plates were left for approximately 30 mins; the antibiotic discs were aseptically transferred directly into the sensitivity plates with the aid of sterile forceps. For gram-positive isolates, ciproflox (CIP, 30 mcg), norfloxacin (NB, 10 mcg), gentamycin (CN, 10 mcg), amoxil (AML, 20 mcg), streptomycin (S, 30 mcg), rifampicin (RD, 20 mcg), erythromycin (E, 30 mcg), chloramphenicol (CH, 30 mcg), ampiclox (APX, 20 mcg) and levofloxacin (LEV, 20 mcg) were used, whereas for gram-negative bacteria septrin (SXT, 30 µg), chloramphenicol (CH, 30 µg), sparfloxacin (SP, 10 µg), ciprofloxacin (CPX, 30 µg), amoxicillin (AM, 30 µg), augmentin (AU, 10 µg), gentamycin (CN, 30 µg), pefloxacin (PEP, 30 µg), tarivid (OFX, 10 µg) and streptomycin (S, 30 µg) were employed. Within 30 minutes of application, the plates were inverted and incubated at 37°C for 18–24 hours.

### Determination of multidrug resistance (MDR)

MDR isolates are those that are resistant to more than two classes of antibiotics tested (Abdu et al., 2018).

### Multidrug-resistance (MDR) index calculation

The MDR index was calculated and interpreted according to the Krumperman (1983) equation: MDR index = a/b, where a = the number of antibiotics to which an isolate was resistant and b = the total number of antibiotics to which the test isolate has been evaluated for sensitivity. A value of the MDR index higher than 0.25 poses a high risk of contamination (Azzam et al., 2017).

### Statistical analysis

Data analysis was performed using SPSS software, version 20. Descriptive statistics were used to analyze the isolates on the basis of frequency and antibiogram patterns.

### Ethical consideration

The ethical approval for this study was obtained from the Cross River State Health Research Ethics Committee (CRS-HREC) with **REC No: CRSMOH/RP/REC/2021/183** and was presented to the administration of UCTH, NNRH and General Hospital Calabar for approval. Before data collection, informed, voluntary, written, and signed consent forms were secured from each study participant. The participants’ data were kept confidential. Positive results were reported to the attending physician for appropriate treatment and management.

## RESULTS

### The prevalence of uropathogens in positive urine samples

In this study, 65 bacterial uropathogens, comprising ten bacterial genera, were isolated from positive urine samples. Among these, gram-negative uropathogens constituted 80% (52/65), whereas 20% (13/65) were gram-positive cocci. The predominant gram-negative bacteria were *Klebsiella pneumoniae* (23.1%), *Escherichia coli* (12.3%), *Enterobacter cloacae* (10.8%), *Citrobacter freundii* (9.2%), *Proteus mirabilis* (7.7%), *Serratia marcescens* (7.7%), *Pseudomonas aeruginosa* (3.1%), *Cronobacter* sp. (3.1%), *Citrobacter koseri* (1.5%) and *Pseudomonas luteola* (1.5%). Among the gram-positive uropathogens, *Staphylococcus* sp. constituted 16.9%, followed by *Enterococcus* sp. (3.1%) (Figure 1).

**Figure 1:**
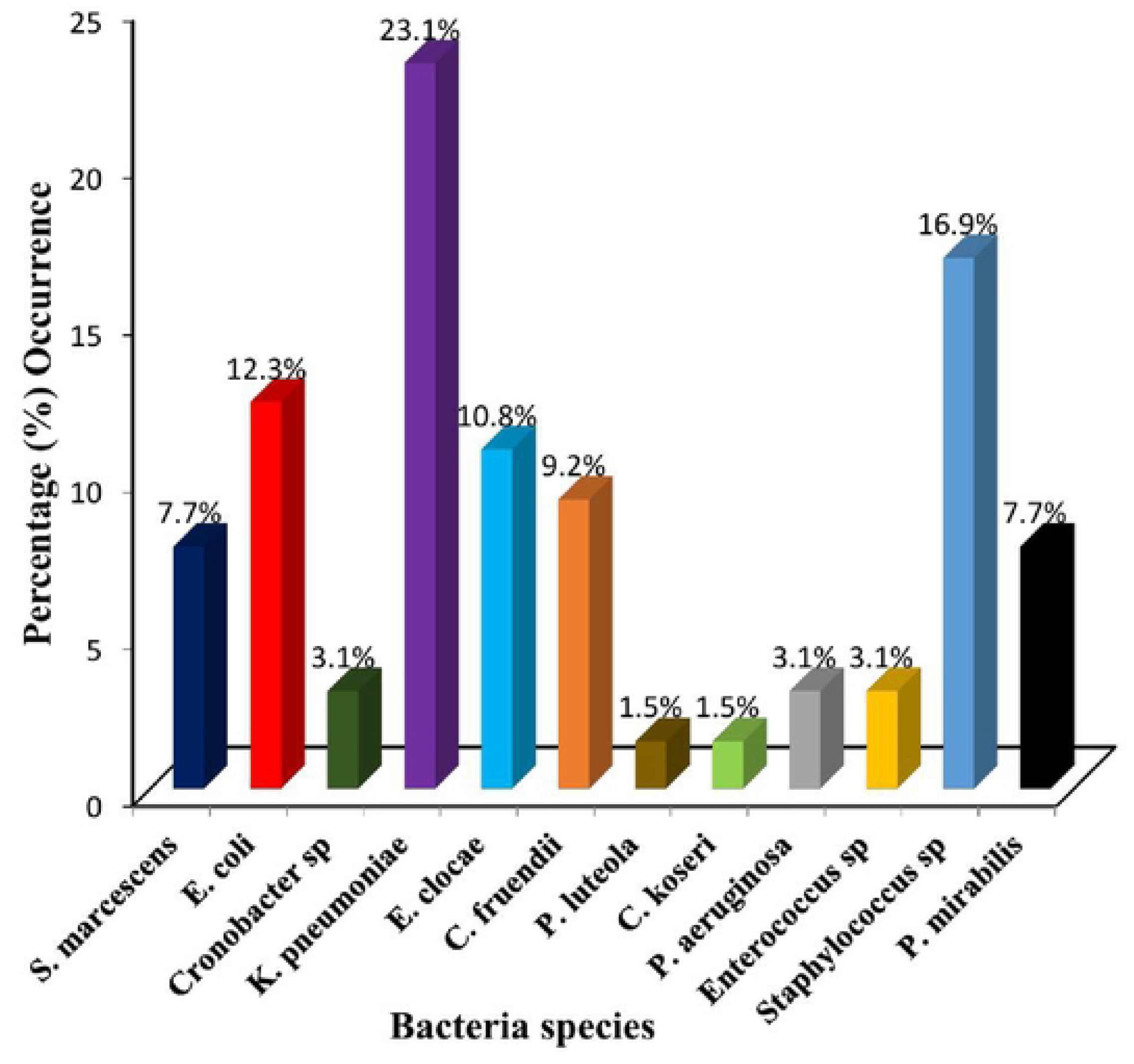
Distribution of bacterial uropathogens isolated from symptomatic UTI patients attending hospitals in Calabar, Nigeria.

### Antibiotic profile of gram-negative uropathogens

Antibiotic susceptibility testing revealed that 98.1% (51/52) of the gram-negative uropathogens isolated from the urine samples were MDR organisms (Table 1). In this study, *S. marcescens* and *E. coli* were completely (100%) resistant to augumentin, septrin, streptomycin and amoxicillin. *E. coli* was 87.5% resistant to both pefloxacin and gentamicin, 62.5% resistant to chloramphenicol and 50.0% resistant to both sparfloxacin and ciprofloxacin. However, *S. marcescens* exhibited 80% susceptibility to sparfloxacin. The antibiotic susceptibility pattern of *Cronobacter* sp. showed 100% resistant to septrin, streptomycin and augmentin. The results revealed that *Cronobacter* sp. was completely (100%) sensitive to sparfloxacin and tarivid, with 50.0% sensitivity to both pefloxacin and gentamicin. *K. pneumoniae* was 100% resistant to augmentin and septrin, 93.3% to amoxicillin, 73.3% to streptomycin, and 60.0% to pefloxacin. The rates of sensitivity of *K. pneumoniae* to sparfloxacin, ciprofloxacin, and gentamicin were 80.0%, 60.0%, and 60.0%, respectively. *Citrobacter* sp. was 100% resistance to amoxicillin, septrin and augmentin and 71.4% resistant to chloramphenicol, tarivid, pefloxacin, gentamicin and streptomycin. The resistance rates of *E. clocae* were 85.7%, 85.7%, 85.7%, 85.7%, 71.4%, 71.4%, 71.4%, 71.4%, 71.4% and 57.1% to augmentin, septin, streptomycin, amoxicillin, chloramphenicol, sparfloxacin, tarivid, pefloxacin, ciprofloxacin and gentamicin, respectively. All *Pseudomonas* sp. strains were completely resistant to amoxicillin, streptomycin, septrin, tarivid, chloramphenicol and augmentin, followed by ciprofloxacin (66.7%). The results revealed that 66.7% of *Pseudomonas* sp. were sensitive to gentamicin. Additionally, *P. mirabilis* was 100% resistance to amoxicillin, septrin and augmentin, followed by pefloxacin (80.0%), gentamicin (80.0%) and 60.0% resistant to sparfloxacin and tarivid. The results revealed that 80.0% of *P. mirabilis* were sensitive to streptomycin (Table 1).

**TABLE 1:** Antibiogram of gram-negative uropathogens isolated from hospitals in Calabar.

| Bacterial uropathogens | N | Pattern | AU | CH | SP | OFX | PEF | CPX | SXT | CN | S | AM |
| --- | --- | --- | --- | --- | --- | --- | --- | --- | --- | --- | --- | --- |
| *S. marcescens* | 5 | R | 5(100) | 2(40) | 1(20) | 2(40) | 3(60) | 0(0.0) | 5(100) | 3(60) | 5(100) | 5(100) |
|  |  | I | 0(0.0) | 1(20) | 0(0.0) | 1(20) | 2(40) | 3(60) | 0(0.0) | 0(0.0) | 0(0.0) | 0(0.0) |
|  |  | S | 0(0.0) | 2(40) | 4(80) | 2(40) | 0(0.0) | 2(40) | 0(0.0) | 2(40) | 0(0.0) | 0(0.0) |
| *E. coli* | 8 | R | 8(100) | 5(62.5) | 4(50) | 5(62.5) | 7(87.5) | 4(50) | 8(100) | 7(87.5) | 8(100) | 8(100) |
|  |  | I | 0(0.0) | 3(37.5) | 2(25) | 2(25) | 0(0.0) | 2(25) | 0(0.0) | 0(0.0) | 0(0.0) | 0(0.0) |
|  |  | S | 0(0.0) | 0(0.0) | 2(25) | 1(12.5) | 1(12.5) | 2(25) | 0(0.0) | 1(12.5) | 0(0.0) | 0(0.0) |
| *Cronobacter* sp | 2 | R | 2(100) | 0(0.0) | 0(0.0) | 0(0.0) | 0(0.0) | 0(0.0) | 2(100) | 0(0.0) | 2(100) | 2(100) |
|  |  | I | 0(0.0) | 2(100) | 0(0.0) | 0(0.0) | 1(50) | 2(100) | 0(0.0) | 1(50) | 0(0.0) | 0(0.0) |
|  |  | S | 0(0.0) | 0(0.0) | 2(100) | 2(100) | 1(50) | 0(0.0) | 0(0.0) | 1(50) | 0(0.0) | 0(0.0) |
| *K. pneumoniae* | 15 | R | 15(100) | 6(40) | 2(13.3) | 8(53.3) | 9(60) | 2(13.3) | 15(100) | 6(40) | 11(73.3) | 14(93.3) |
|  |  | I | 0(0.0) | 4(26.7) | 1(6.7) | 0(0.0) | 1(6.7) | 4(26.7) | 0(0.0) | 0(0.0) | 1(6.7) | 1(6.7) |
|  |  | S | 0(0.0) | 5(33.3) | 12(80) | 7(46.7) | 5(33.3) | 9(60) | 0(0.0) | 9(60) | 3(20) | 0(0.0) |
| *E. cloacae* | 7 | R | 6(85.7) | 5(71.4) | 5(71.4) | 5(71.4) | 5(71.4) | 5(71.4) | 6(85.7) | 4(57.1) | 6(85.7) | 7(100) |
|  |  | I | 0(0.0) | 1(14.3) | 0(0.0) | 1(14.3) | 1(14.3) | 0(0.0) | 0(0.0) | 1(14.3) | 0(0.0) | 0(0.0) |
|  |  | S | 1(14.3) | 1(14.3) | 2(28.6) | 1(14.3) | 1(14.3) | 2(28.6) | 1(14.3) | 2(28.6) | 1(14.3) | 0(0.0) |
| *Citrobacter* sp | 7 | R | 7(100) | 5(71.4) | 4(57.1) | 5(71.4) | 5(71.4) | 3(42.9) | 7(100) | 5(71.4) | 5(71.4) | 7(100) |
|  |  | I | 0(0.0) | 2(28.6) | 1(14.3) | 1(14.3) | 2(28.6) | 2(28.6) | 0(0.0) | 0(0.0) | 1(14.3) | 0(0.0) |
|  |  | S | 0(0.0) | 0(0.0) | 2(28.6) | 1(14.3) | 0(0.0) | 2(28.6) | 0(0.0) | 2(28.6) | 1(14.3) | 0(0.0) |
| *Pseudomonas* sp | 3 | R | 3(100) | 3(100) | 1(33.3) | 3(100) | 1(33.3) | 2(66.7) | 3(100) | 1(33.3) | 3(100) | 3(100) |
|  |  | I | 0(0.0) | 0(0.0) | 1(33.3) | 0(0.0) | 2(66.7) | 0(0.0) | 0(0.0) | 0(0.0) | 0(0.0) | 0(0.0) |
|  |  | S | 0(0.0) | 0(0.0) | 1(33.3) | 0(0.0) | 0(0.0) | 1(33.3) | 0(0.0) | 2(66.7) | 0(0.0) | 0(0.0) |
| *P. mirabilis* | 5 | R | 5(100) | 2(40) | 3(60) | 3(60) | 4(80) | 2(40) | 5(100) | 4(80) | 1(20) | 5(100) |
|  |  | I | 0(0.0) | 3(60) | 1(20) | 2(40) | 1(20) | 3(60) | 0(0.0) | 0(0.0) | 0(0.0) | 0(0.0) |
|  |  | S | 0(0.0) | 0(0.0) | 1(20) | 0(0.0) | 0(0.0) | 0(0.0) | 0(0.0) | 1(20) | 4(80) | 0(0.0) |
| Total n (%) | 52 | **R** | **51(98.1)** | **28(53.8)** | **20(38.5)** | **31(59.6)** | **34(65.4)** | **18(34.6)** | **51(98.1)** | **30(57.7)** | **41(78.8)** | **48(92.3)** |
|  |  | I | 0(0.0) | 16(30.8) | 6(11.5) | 7(13.5) | 10(19.2) | 16(30.8) | 0(0.0) | 2(3.8) | 2(3.8) | 3(5.8) |
|  |  | **S** | **1(1.9)** | **8(15.4)** | **26(50)** | **14(26.9)** | **8(15.4)** | **18(34.6)** | **1(1.9)** | **20(38.5)** | **9(17.3)** | **1(1.9)** |

R: Resistant; I: Intermediate; S: Susceptible; N: Number of isolates; AU: Augmentin; CH: Chloramphenicol; SP: Sparfloxacin; OFX: Tarivid; PEF: Pefloxacin; CPX: Ciprofloxacin; SXT: Septrin; CN: Gentamycin; S: Streptomycin; AM: Amoxicillin

In general, 98.1% of the gram-negative uropathogens were resistant to β-lactam (augmentin) or sulfonamide (septrin), followed by amoxacillin (92.3%) and streptomycin (78.8%). However, ciprofloxacin and sparfloxacin were the most effective drugs against gram-negative uropathogens, with overall resistance rates of 34.6% and 38.5%, respectively (Figure 2). The gram-negative uropathogens were resistance to 3-10 antibiotics, and twenty-four different phenotypic resistance profiles were observed (Table 2). The multiple antibiotic resistance index (MDR-I) of gram-negative uropathogens ranges from 0.3-1.0. The highest MDR-I values observed in this study were 0.6 and 1.0, with 21.6% each. Seven (13.7%), 6 (11.8%), 5 (9.8%), 4 (7.8%) and 3 (5.9%) isolates had MDR-I values of 0.4, 0.8, 0.7, 0.5, and 0.9, and 0.3 respectively (Table 2).

**Figure 2:**
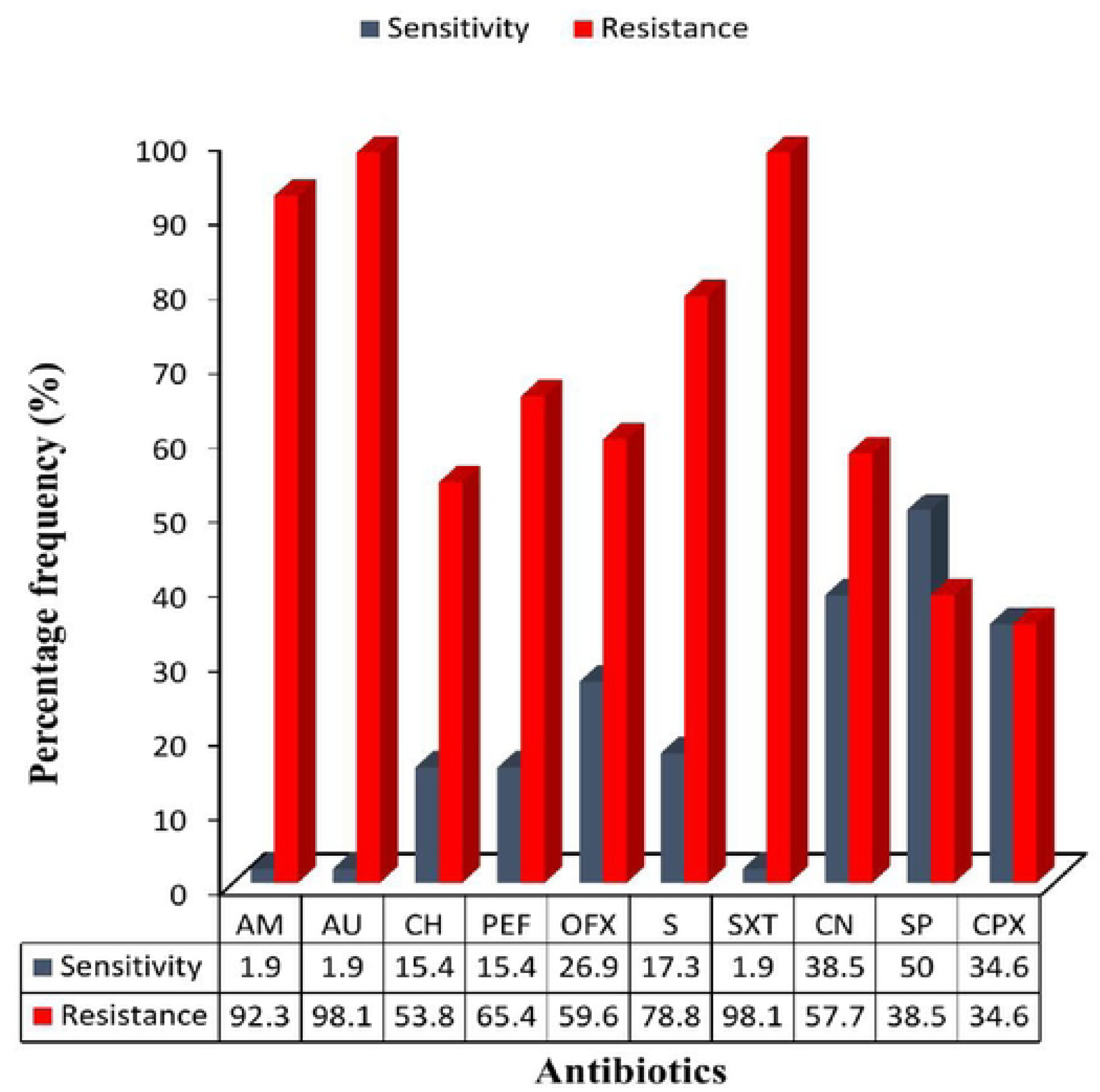
Overall percentage susceptibility and resistance of gram-negative uropathogens KEY: AM: Amoxicillin; AU: Augmentin; CH: Chloramphenicol; PEF: Pefloxacin; OFX: Tarivid; S: Streptomycin; SXT: Septrin; CN: Gentamycin; SP: Sparfloxacin; CPX: Ciprofloxacin.

**TABLE 2:**
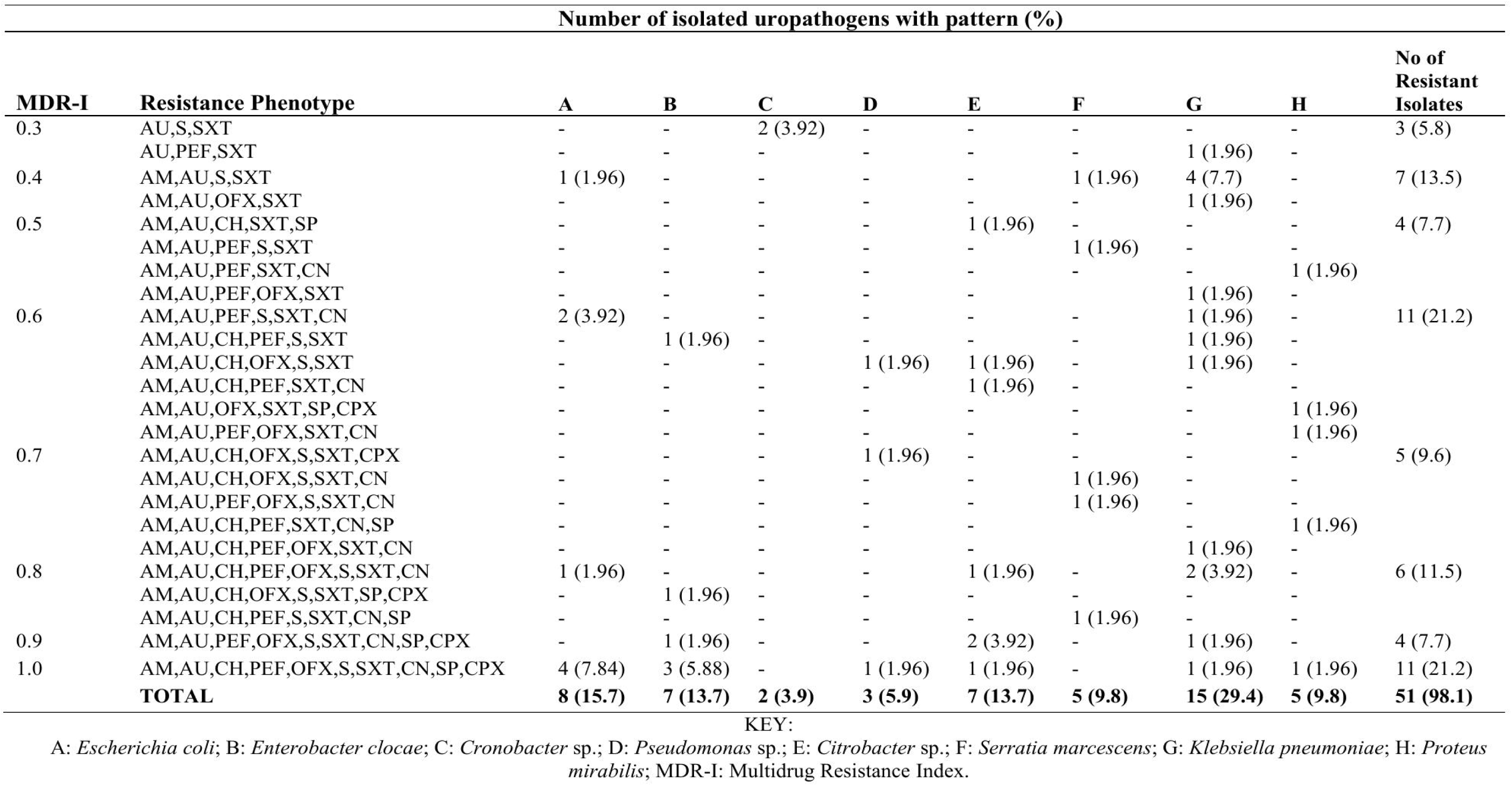
Multidrug resistance (MDR) phenotypes of gram-negative uropathogens from hospitals in Calabar.

### Antibiotic profile of gram-positive uropathogens

Among the gram-positive uropathogens tested, *Staphylococcus* sp. was 100% resistant to amoxil, erythromycin, ampiclox and rifampicin (Table 3). On the other hand, the susceptibilities of *Staphylococcus* sp. to the tested antibiotics were 81.8%, 63.6%, and 54.5% for levofloxacin, ciprofloxacin, and streptomycin, respectively (Table 3). *Enterococcus* sp. showed very high (100%) resistance to norfloxacin, gentamycin, amoxil, ampiclox, erythromycin and rifampicin. The most effective drugs against *Enterococcus* sp. were chloramphenicol and levofloxacin, showing 100% susceptibility (Table 3). All gram-positive uropathogens isolated in this study were MDR. The gram-positive uropathogens were resistant to 4-9 antibiotics, and nine different types of phenotypic resistance profiles were observed (Table 4). As shown in Table 4, two isolates were resistant to 4 antibiotics, four were resistant to 5 antibiotics, three were resistant to 6 antibiotics, two were resistant to 7 antibiotics, and one was resistant to 7 or 8 antibiotics. The MDR-I values of gram-positive uropathogens ranged from 0.4-0.9. The highest MDR-I observed in this study was 0.5, with a value of 30.8%. Three (23.1%) had MDR-I values of 0.6, 2 (15.4%) had MDR-I values of 0.4 and 0.7 each, and 1 (7.7%) had MDR-I value of 0.8 and 0.9 each (Table 4). The overall results indicate that fluoroquinolone, chloramphicol and streptomycin were the most active drugs against gram-positive uropathogens (Figure 3).

**Figure 3:**
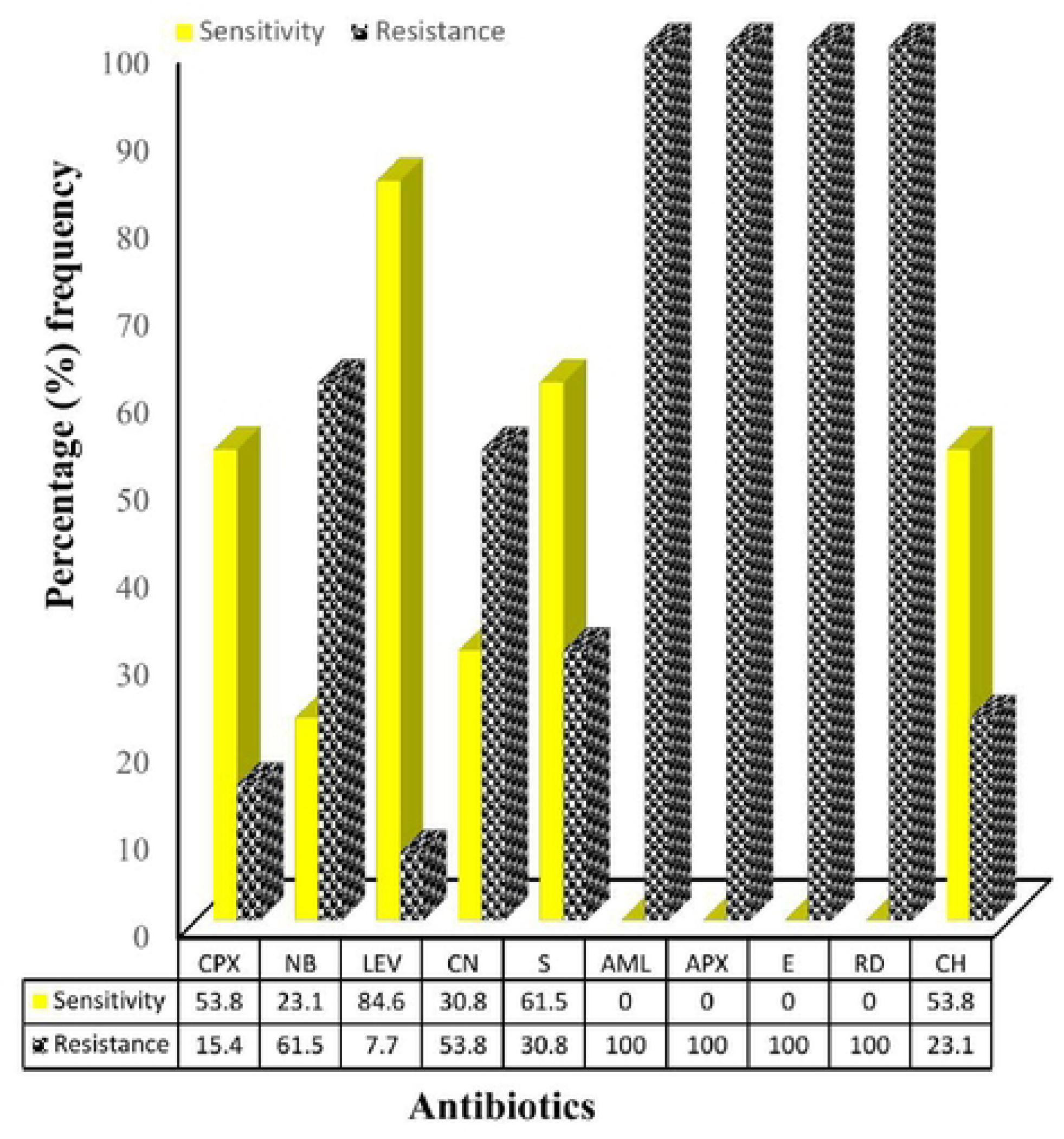
Overall percentage susceptibility and resistance of gram-positive uropathogens KEY: CPX: Ciproflox; NB: Norfloxacin; LEV: Levofloxacin; CN: Gentamycin; S: Streptomycin: AML: Amoxil; APX: Ampiclox; E: Erythromycin; RD: Rifampicin; CH: Chloramphenicol.

**TABLE 3:**
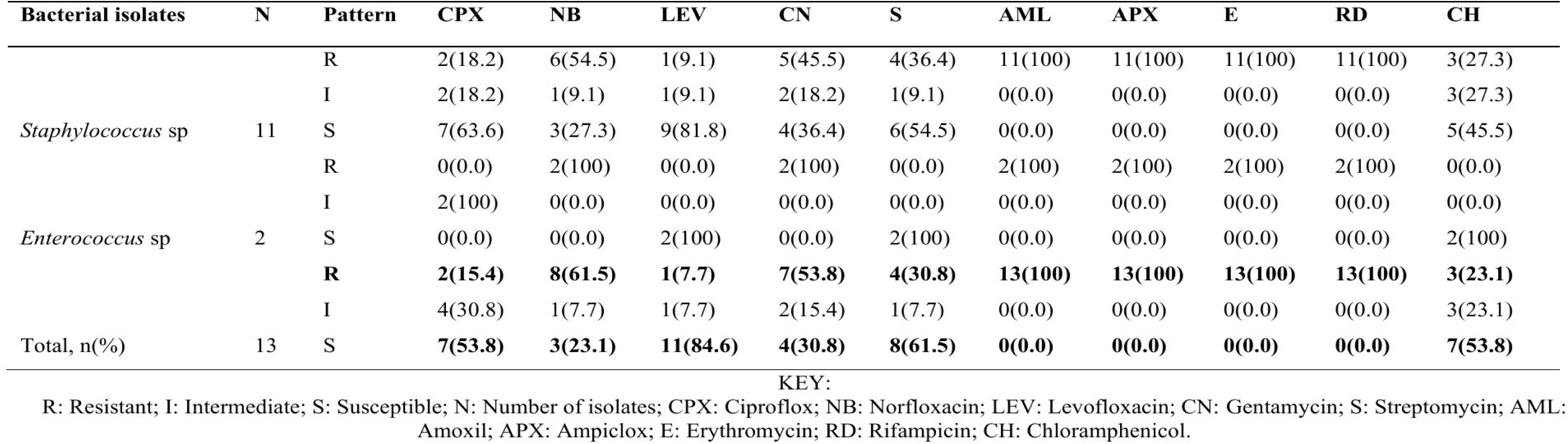
Antibiogram of gram-positive uropathogens isolated from hospitals in Calabar.

**TABLE 4:**
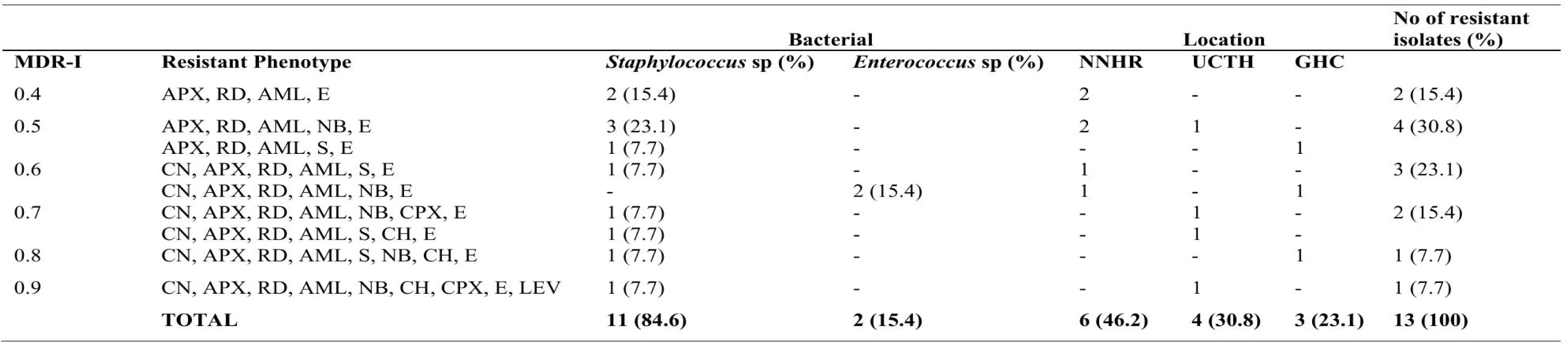
Multidrug resistance (MDR) phenotypes of gram-positive uropathogens from hospitals in Calabar.

## DISCUSSION

The etiology and susceptibility of uropathogens to available antibiotics varies with location (Bassey et al., 2024). To reduce the burden of the disease, adequate knowledge of the etiology and antibiogram of uropathogens in a particular setting is crucial for effective treatment. This study aimed to determine the status of antibiotic resistance among bacterial uropathogens from symptomatic patients attending tertiary healthcare facilities in Calabar, southern Nigeria. In our study, *K. pneumoniae* was the most common uropathogen (23.1%), followed by *Staphylococcus* sp. (16.9%) and *Escherichia coli* (12.3%). The high prevalence of *K. pneumoniae* in this study is an indication that the organism is becoming more prominent as a causative agent of UTI. Similarly, Mistry et al. (2022) reported *K. pneumoniae* (31.37%) as the most predominant uropathogen in Rajkot, India. Additionally, Vinod and Selvaraj (2012) reported *K. pneumoniae* (65%) as the most common uropathogen among the Paliyars tribe living in Tamil Nadu, India. However, our study contradicts the studies of Seifu and Gebissa (2018) and Alo et al. (2015), who reported *E. coli* as a common uropathogen from UTI patients.

In other studies, Many et al. (2019) reported *P. mirabilis* (41.2%) in the DRC, Labi et al. (2015) reported *Enterococcus* sp (26.7%) in Ghana and Musonda et al. (2020) reported *Staphylococcus aureus* (32%) in Zambia. The variation in the type of bacterial uropathogens in this study and other studies might be attributed to the physiological state of patients, techniques of sample collection, sample size used, and environmental or personal hygiene (Seifu & Gebissa, 2018; Vinod & Selvaraj, 2012). In this study, gram-negative uropathogens constituted 80% and gram-positive pathogens constituted 20%. These findings support the assertion that gram-negative bacteria constitute 80–90% of uropathogens (Vinod & Selvaraj, 2012). The results of the present study are in line with those of Alo et al. (2015) and Khoshbakht et al. (2013), who reported that *Staphylococcus* sp. was the second most prevalent bacterial uropathogens associated with UTI.

Another unique observation in this study was the presence of *Cronobacter* sp. (3.1%) in the analyzed urine samples. *Cronobacter* sp. have been reported as emerging pathogens from infant food (Pakbin et al., 2022). The incidence of UTI caused by *Cronobacter* sp. in the study area is rear. However, this study corroborated the findings of Hayashi et al. (2021), who reported the occurrence of *C. sakazakii* in a 69-year-old man who presented with a UTI at Shimane University Hospital, Shimane, Japan. The possible route of infection with *Cronobacter* sp. could be via oral ingestion from external sources and retrograde UTI (Hayashi et al., 2021).

The emergence of bacteria resistant to available antibacterial products remains a global concern in disease management. This current study revealed that 98.1% of the gram-negative uropathogens recovered from UTI patients demonstrated MDR to each of augmentin and septrin, followed by amoxicillin (92.3%) and streptomycin (78.8%). High resistance of gram-negative uropathogens to these classes of antibiotics has been documented in several studies (Marami et al., 2019; Dibua et al., 2014; Iheanacho et al., 2018). The observed resistance rate is worrisome in that the tested antibiotics are among the commonly prescribed drugs for the treatment of UTIs (Terlizzi et al., 2017). These results are similar to those of Worku et al. (2021), who reported 100% MDR among gram-negative uropathogens in Addis Ababa, Ethiopia. Additionally, the observed resistance among gram-negative uropathogens is comparable to the 87.2% reported by Gebremariam et al. (2019). The decline in the activity of amoxicillin, augmentin, septrin and streptomycin in the present study may be due to the abuse of these drugs within the study area. In addition, gram-negative bacteria, particularly *K. pneumoniae*, *E. coli*, *P. mirabilis* and *P. aeruginosa,* are known to produce extended-spectrum β-lactamases (ESBLs). The presence of ESBL genes in the isolated uropathogens may have contributed to the observed MDR (Giwa et al., 2018; Belete, 2020; Chaula et al., 2016). However, among the antibiotics tested, gentamicin and sparfloxacin were the most effective drugs against gram-negative uropathogens.

In contrast to the previously reported susceptibility rates of gram-negative uropathogens to ciprofloxacin (77.6%) reported in Akure, Nigeria (Simon-Oke et al., 2019) and Harar, in Ethiopia (74.1%) (Marami et al., 2019), a lower susceptibility rate was observed in this study (34.6%), comparable to the work of Worku et al. (2021), who reported a 38.9% susceptibility rate in Addis Ababa, Ethiopia. The difference in the susceptibility rate could be affected by the geographical distribution and variability of strains among the same species.

Gram-positive bacteria detected in this study were highly resistant to amoxil, ampiclox, erythromycin, and rifampicin (100%), indicating that resistance to these drugs is widespread in Calabar. This resistance rate was similar to that reported in Enugu by Dibua et al. (2014), who reported 100% resistance to β-lactam drugs. Also, the ineffectiveness of rifampicin against gram-positive uropathogens in this study is comparable to that reported by Mistry et al. (2022) in Rajkot, India. The high resistance rate could be attributed to the production of penicillinase enzymes, indiscriminate self-administration of drugs by patients and other anthropogenic factors, which help organisms become resistant to β-lactam antibiotics (Gebremariam et al., 2019). In general, gram-positive uropathogens were more susceptible to levofloxacin (84.6%), streptomycin (61.5%), ciprofloxacin and chloramphenicol (53.8%) each. This is true considering previous reports by Gebremariam et al. (2019) in Ethiopia and Simon-Oke et al. (2019) in Akure, Nigeria, where fluroquinolone, chloramphenicol and streptomycin were found to be effective against gram-positive uropathogens. Antibiotic resistance among uropathogens is clearly increasing. The abuse of antibiotics and lack of awareness of the dangers of antibiotic misuse are considered the most important factors in the spread of antibiotic resistance in the community (Kourkouta et al., 2017). There is no doubt that the world is in the cusp of post-antibiotic era and alternative to antibiotics are urgently needed for the treatment of bacterial infections (Mohammed et al., 2023), and phage therapy is a promising approach to combat antibiotic-resistant infections (Mohammed & Orzechowska, 2021).

## CONCLUSION

Urinary tract infections (UTIs) are major health concern affecting millions of people globally each year, and their management is complicated by increasing resistance among uropathogens to antibiotics, leading to increased morbidity and healthcare costs. In many settings, especially in developing countries, empiric antibiotic administration is recognized as a contributory factor exacerbating drug resistance. In this study, *K. pneumoniae* was the predominant uropathogens, followed by *Staphylococcus* sp. Other isolates recovered were *E. coli*, *E. clocae*, *C. freundii*, *P. mirabilis*, *S. marcescens*, *P. aeruginosa*, *Cronobacter* sp., *Enterococcus* sp., *C. koseri*, and *P. luteola*. Most of the isolates were highly resistant to augmentin, amoxicillin, septrin, ampicillin, erythromycin, and rifampicin. MDR was observed in 98.1% of the gram-negative isolates, whereas 100% of the gram-positive uropathogens were MDR. However, levofloxacin, gentamicin, ciprofloxacin and sparfloxacin were effective against the isolates. This study revealed a high rate of MDR index (˃0.25) for commonly prescribed antibiotics. To reduce the emergence of resistance in bacterial uropathogens, prudent use of antimicrobial agents is advised.

## Data Availability

All data produced in the present work are contained in the manuscript

## Notes

### Competing Interest Statement

The authors have declared no competing interest.

### Author Declarations

The ethical approval for this study was obtained from the Cross River State Health Research Ethics Committee (CRS-HREC) with REC No: CRSMOH/RP/REC/2021/183 and was presented to the administration of UCTH, NNRH and General Hospital Calabar for approval.

